# Test-retest reliability of hepatic venous pressure gradient and impact on trial design: a study in 289 patients from the control groups of 20 randomized trials

**DOI:** 10.1101/2020.12.07.20245464

**Authors:** Wayne Bai, Mustafa Al-Karaghouli, Jesse Stach, Granville J Matheson, Juan G Abraldes

## Abstract

**Background and Aims:** Portal hypertension (PH) is a major driver for cirrhosis complications. Portal pressure is estimated in practice by the hepatic venous pressure gradient (HVPG). The assessment of HVPG changes has been used for drug development in PH. This study aimed at quantifying the test-retest reliability and consistency of HVPG in the specific context of RCTs for the treatment of PH in cirrhosis and its impact on power calculations for trial design.

**Method:** We conducted a search of published RCTs in patients with cirrhosis reporting individual patient-level data of HVPG at baseline and after an intervention, and that included a placebo or untreated control arm. Baseline and follow-up HVPG in the control groups were extracted after digitizing the plots. We assessed different reliability parameters and the potential impact of study characteristics.

**Results:** We retrieved a total of 289 before-after HVPG measurements in the placebo/untreated groups from 20 RCTs. Time range between the two HVPGs measurements was 20 min to 730 days. Test-retest reliability was higher in studies including only compensated patients and, therefore, modelled sample size calculations for trials in compensated cirrhosis were lower than for decompensated cirrhosis. Higher proportion of alcohol-related cirrhosis and unicentric trials were associated with lower differences between baseline and follow-up measurements. Smallest detectable difference in an individual was 24% and 32% in compensated and decompensated patients respectively

**Conclusion:** The test-retest reliability of HVPG is overall excellent, but higher in studies limited to compensated cirrhosis. These findings should be taking into account when powering trials based in the effects on HVPG or when consider HVPG as a tool to guide therapy of portal hypertension

## Introduction

Chronic liver disease (CLD) and cirrhosis causes major mortality and morbidity worldwide accounting for 2 million deaths internationally each year^1^. The main factor determining liver-related mortality in cirrhosis is the development of decompensation^2^. A major factor driving the development of decompensation is the progressive increase in portal pressure, and for every increase in one mmHg in portal pressure gradient there is a 11% relative increase in the risk of decompensation^3,4^. Furthermore, a very recent randomized trial showed that, in compensated patients, decreasing portal pressure with beta-adrenergic blockers decreases the risk of cirrhosis decompensation^5^.

In clinical practice hepatic venous pressure gradient (HVPG) is the gold standard for determining the portal pressure ^6^. This is measured by occluding one of the hepatic veins either with a balloon tipped catheter or with a straight catheter advanced until getting it wedged in the hepatic vein ^7^. The HVPG is calculated as the difference between the wedged or occluded pressure, and the free pressure in the hepatic vein which acts as an internal zero ^8,9^. Several technical reviews and a renewed interest in its use for drug development in NAFLD have helped with standardization and quality assurance of HVPG measurements^4,10-12^.

Several studies have consistently shown an association between the changes in HVPG and clinically relevant outcomes. On that basis, the assessment of HVPG changes has been the gold standard for the drug development for portal hypertension^13,14^. Moreover, there is increasing interest in wider use of HVPG as a clinical outcome measure in RCTs for the assessment of etiological treatments of cirrhosis^15^.

The use of HVPG as a prognostic indicator or as an outcome to assess the response to a drug, either in a clinical context or in the context of drug development, requires a detailed understanding of the reliability of the measurements. Reliability relates the proportion of the total variance which is not attributable to error, thereby describing the ability of a measure to distinguish between individuals. Variation in HVPG measurements might be related to circadian rhythms ^16^, use or anaesthesia or sedation ^17^, changes in the disease trajectory (improving or worsening) that have an impact on the degree of portal hypertension ^18-20^ or measurement errors. For example, the presence of veno-venous communications introduces a systematic error, since it results in lower HVPG than true portal pressure gradient, and therefore measures with these characteristics should be deemed unreliable^21^. On the other hand, it has been shown that technical choices, such as the use of a straight catheter rather than a balloon catheter, can increase the random error in HVPG measurement ^22^. Finally, reading of permanent HVPG tracings can introduce further random error, though this is associated with minimal interobserver variability in experienced hands ^23^, which has led to recent introduction of centralized expert reading in RCTs using HVPG ^4,10,12^

The design of therapeutic trials using measurements of HVPG have paid little attention to its measurement error. The reliability and consistency of HVPG measurements will impact sample size and power calculations ^24^, since with higher levels of measurement error, a study has lower power to detect a given underlying effect size. A greater understanding of the measurement properties of HVPG is therefore of importance to assess the feasibility of new studies in which HVPG is the main outcome.

Therefore, the aims of this study were to quantify the reliability and consistency of HVPG measurement, to understand potential factors which influence its measurement, and provide guidelines for power analysis in future studies. Since our major interest was to assess the impact of HVPG measurement error on the design of trials for drug development of portal hypertension, we conducted our study in this particular context, using previously published individual patient data from the placebo or untreated arms of 20 RCTs aimed at evaluating the effect of a drug on portal hypertension.

## Methods

We included published RCTs in patients with cirrhosis with published individual patient-level HVPG data, before and after the intervention, and that included a placebo or untreated control arm.

### Search strategy and data extraction

This is thoroughly described in supplementary material S1

### Outcome Measures

The test-retest reliability measures were assessed by the intraclass correlation coefficient for absolute agreement (ICC) and by the smallest detectable difference (SDD)^24^. The ICC within each study reflects the proportion of the variance explained by the grouping structure within that study, which in our case was the individual patient (each patient contributing with two measurements). Thus, if all patients had identical pre-post HVPG, the ICC would have a value of 1. Since the variance of HVPG within an individual is scaled to the variance of HVPG within the study population, for a given magnitude of within-individual variability, the wider the distribution of HVPG between the individuals of the study, the higher would be the ICC. This scaling reflects the fact that a small degree of error can be problematic for differentiating individuals when they differ very little, or that relatively large errors can be acceptable when individuals differ substantially. A figure to illustrate this concept is provided as supplementary materials S2. We considered ICC>0.75 as good, and above 0.9 as excellent and acceptable for diagnostic purposes^25^. An ICC between 0.50 and 0.75 was considered poor to moderate reliability.

The SDD (also referred to as the minimum detectable difference) between two measurements *in a given subject* refers to the difference which would be sufficiently large in an individual patient as to be considered unlikely to have been due to chance alone, according to a 95% confidence interval.

To assess potential modifiers of the reliability of HVPG in clinical trials we collected several study characteristics, including the proportion of patients with decompensated cirrhosis, time between measurements, type of catheter used for HVPG measurements (balloon vs wedged catheter) and proportion of patients with alcohol-related liver disease. Even if we had the individual HVPG data, in most studies it was not possible to assign individual patient characteristics to individual HVPG measurements. Since it has been recommended to conduct studies for compensated and decompensated patients separately^14^, we used this study characteristic as the main one to provide metrics of reliability and consistency, and to define sample size calculation. The studies by Abraldes^26^ and Garcia-Tsao^11^ provided separate data for compensated and decompensated patients, and were thus analysed as two different studies. For the rest of the studies, we analysed those variables as study-level characteristics.

### Statistical analysis

Details of statistical analysis are provided in Supplementary data S3. All raw data, analysis code and additional figures can be found at the following link: https://github.com/mathesong/HVPG_TRT

## Results

### Study selection and study characteristics

We initially identified 689 manuscripts out of which 20 met the inclusion criteria^11,26-29 30-32 33-36 37-40 41-44^ The main characteristics of the 20 RCTs are summarized in table 1 and supplementary materials S4.

**Table 1.**
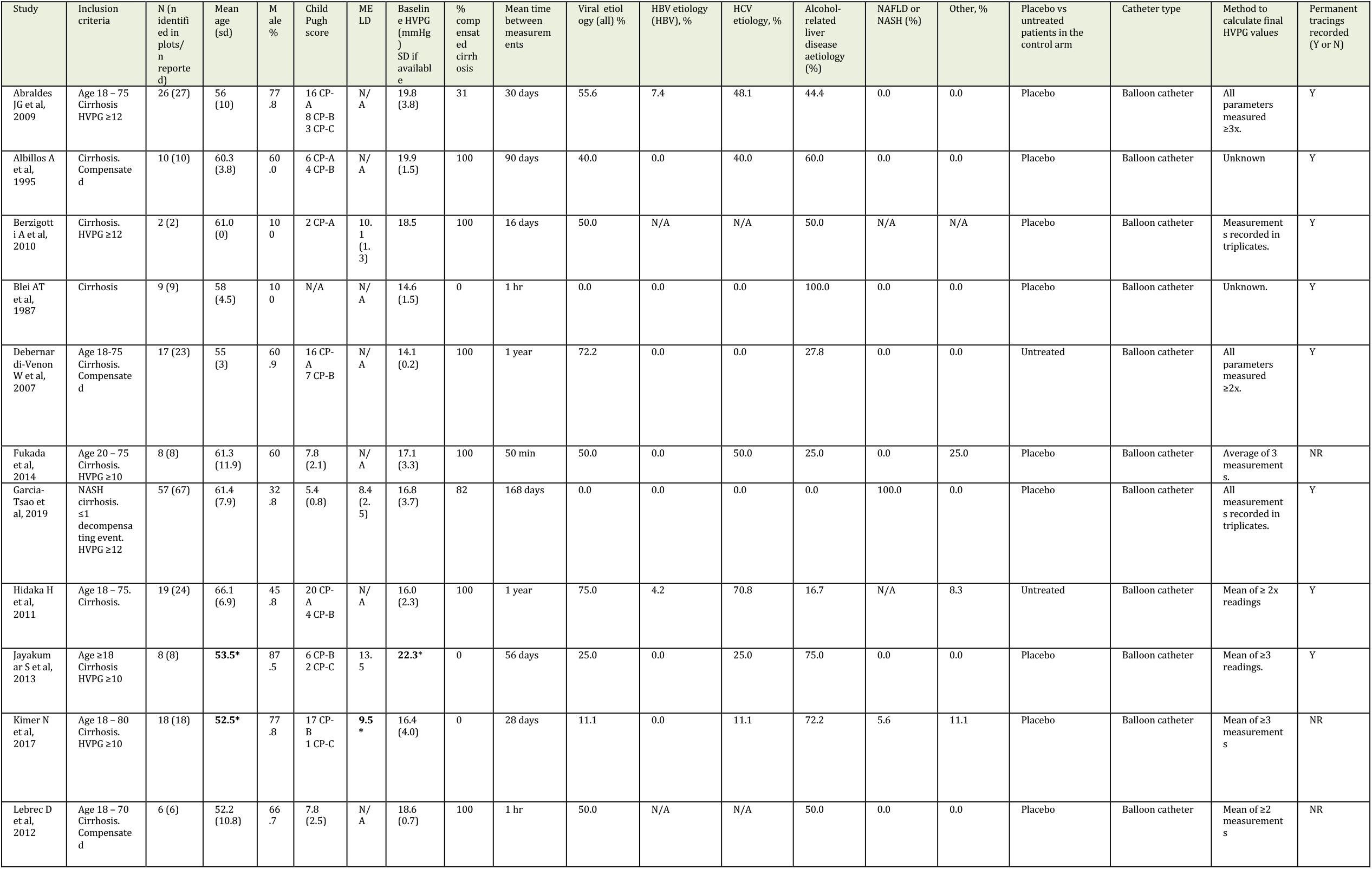
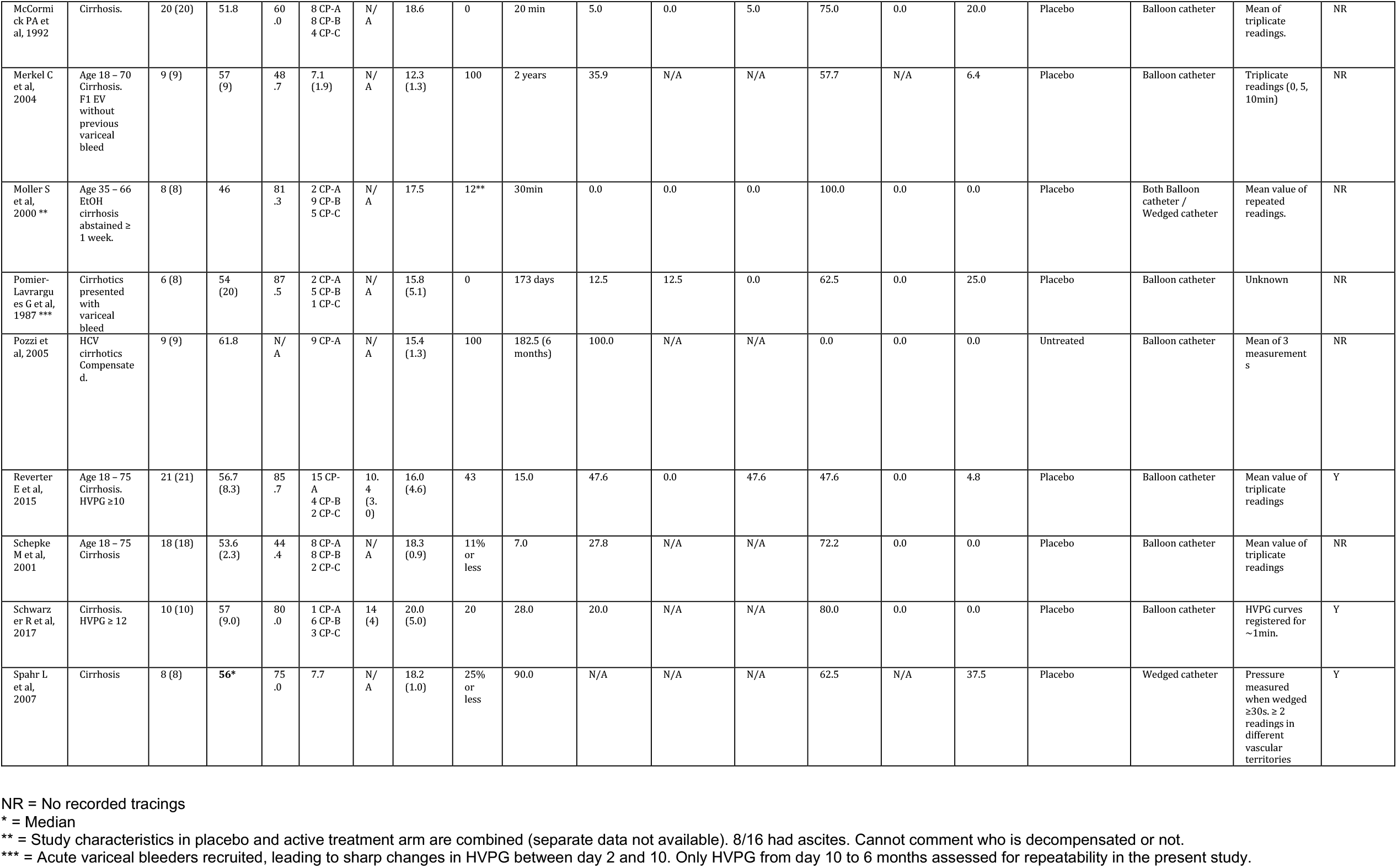
Main characteristics of Included studies.

### Test-retest reliability of HVPG

Metrics of dispersion and test-retest variability for studies including and not including decompensated cirrhosis are shown in Table 2. Supplementary materials S5 shows these metrics for each of the included studies, together with study characteristics. Figure 1a shows individual ICCs. The only study performed exclusively with the straight wedged catheter^34^ was the one with lowest ICC, and was removed for subsequent power calculations, since it is well established that the straight catheter provides more variable results^22^, and the balloon catheter has become the standard for randomized trials. The pooled ICC was numerically higher (better) for studies including compensated patients (0.87, 95% CI: 0.84 - 0.90) than for studies including decompensated patients (0.82, 95% CI: 0.77 - 0.86), though this difference was not significant (one-sided bootstrap test of difference in ICC: p=0.120; difference 95% CI: -0.03 - 0.15).

**Table 2:** Test-retest metrics for the total sample for both patient groups.

| **Patients** | **Mean**<br>**HVPG** | **CV** | **WSCV** | **ICC** | **SDD** | **Change SD** |
| --- | --- | --- | --- | --- | --- | --- |
| Only<br>Compensated | 16.2 | 0.24 | 0.09 | 0.87 | 3.92 | 2.01 |
| Includes<br>Decompensated | 17.79 | 0.27 | 0.12 | 0.82 | 5.68 | 2.91 |

CV: coefficient of variation. WSCV: Within-subject coefficient of variation. ICC: intraclass correlation coefficient. SDD: smallest detectable difference. Change SD: the standard deviation of the signed change between measurements.

**Fig 1:**
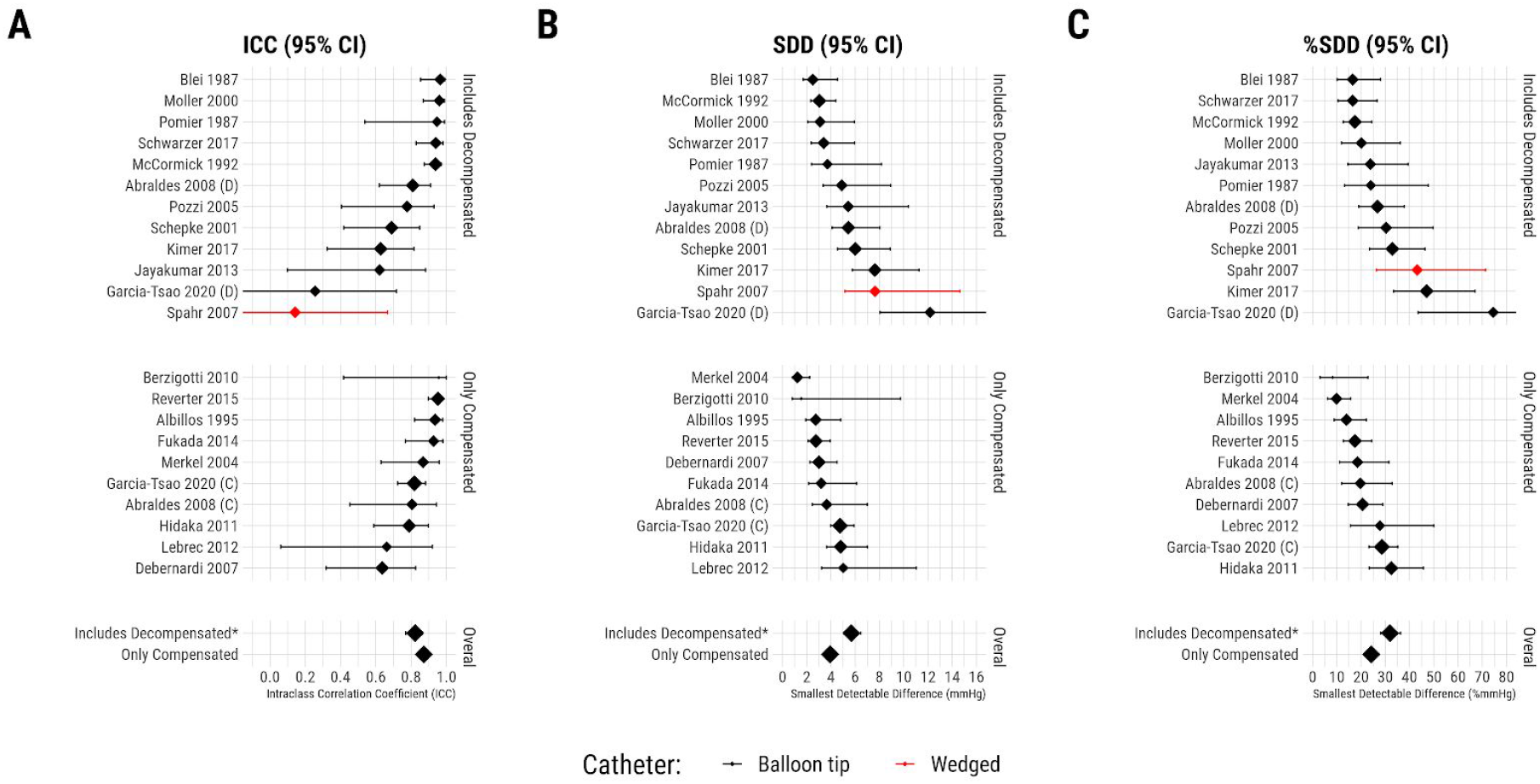
Forest plot showing Intraclass correlation coefficients (ICC) (A), smallest detectable differences (SDD) in mmHg (B), and SDD expressed in % (C), where the size of the point represents the sample size. The studies with a (C) and (D) represent the subsamples of these studies with compensated and decompensated patients respectively. The overall values are presented below, and * depicts that this refers only to studies conducted using balloon-tip catheters. This plot shows that the ranking of the different studies is not the same, since ICC and SDD reflect reliability and consistency respectively. ICC estimates reliability, a measure of differentiability, which is primarily important in between-subjects designs, while the SDD is a measure of consistency, which is primarily important in within-subjects designs.

Figure 1b shows the smallest detectable differences (SDDs) for the different studies. Studies performed in compensated cirrhosis showed significantly (one-sided bootstrap test of difference in ICC: p=0.002; difference 95% CI: 0.56 - 2.97) lower (better) pooled SDD (3.9 mmHg, 95% CI: 3.5 - 4.4) than studies including decompensated patients (5.7 mmHg, 95% CI: 5.1 - 6.5). This was also the case when considering SDD in % change: pooled SDD was 24% (95% CI: 22% to 27%) for studies in compensated patients, and 32% (95% CI: 28% to 36%) in studies including decompensated patients (one-sided bootstrap test of difference in ICC: p=0.015; difference 95% CI: 0.3% - 14.8%) (fig 1c).

### Individual changes in HVPG and potential modifiers

#### Mean values

Figure 2A and 2B show the distribution of individual changes in HVPG in patients from studies with compensated patients and in those with decompensated patients. The median difference between test and retest was 0 mmHg, both in the total sample, as well as in both subsamples of studies containing only compensated cirrhosis patients as well as those including decompensated patients, and did not significantly differ from zero in either the total sample (intercept = -0.037, t_9.5_=-0.189, p=0.85), or in either the compensated (intercept = -0.01, t_6.35_=-0.064, p=0.95) or decompensated (intercept = -0.05, t_5.60_=-0.064, p=0.89) patient groups. Mean values were not significantly associated with absolute change values (Total: estimate = mmHg, p=0.384; Compensated: r=0.065, p=0.396; Includes decompensated: r = 0.05, p = 0.6), however they were significantly associated with signed change values: higher mean values were associated with increases, while lower mean values associated with decreases in retest values compared to baseline (Total: estimate = 0.12 mmHg, t_267.0_ = 3.5, p < 0.001; Compensated: estimate = 0.13, t_134.9_ = 2.98, p = 0.003; Includes decompensated: estimate = 0.12, t_127.5_ = 2.2, p = 0.03).

**Fig 2:**
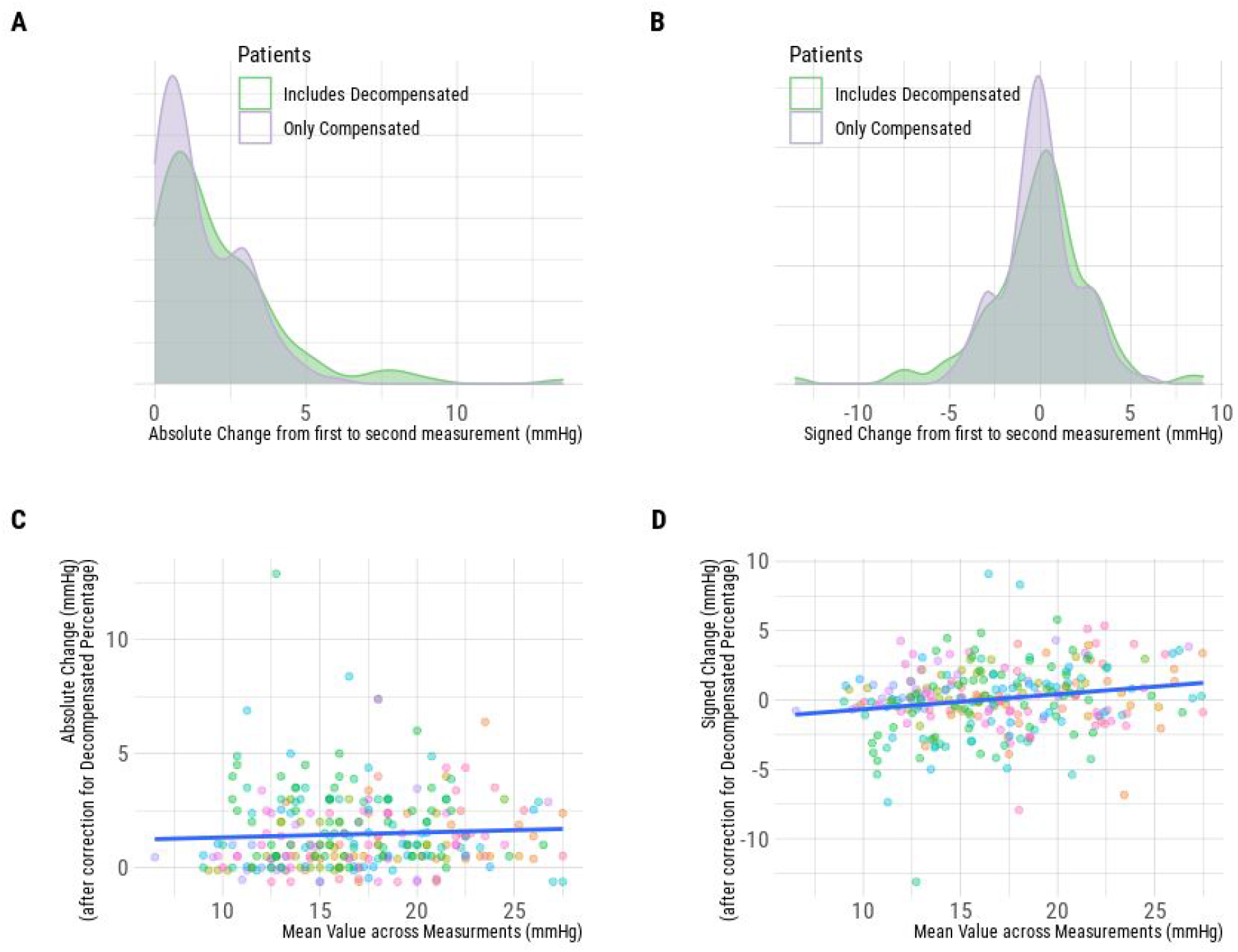
Absolute and signed changes between measurements in relation to the mean value across measurements. (A) Distribution of absolute changes in mmHg divided by patient groups. Note that there is a higher proportion of decompensated patients with higher absolute changes. (B) Distribution of signed changes in mmHg divided by patient groups. Note that both groups are centred around zero. The association between mean values and mean values (C) and signed values (D) after correction for decompensated percentage. For absolute values, this association was not significant, but for signed values, this association was significant in both groups and in total. Colours represent different studies.

#### Study characteristics

We then assessed which study characteristics had an impact on HVPG measurement consistency by assessing their association with the absolute differences between baseline and followup. Studies including higher proportions of decompensated cirrhosis patients had higher pre-post HVPG variability than studies including only compensated cirrhosis (r = 0.18, p= 0.002, estimate = 0.007 mmHg between 0% and 100%, figs 3A). We assessed the remaining characteristics after correction for the proportion of decompensated patients. The average number of days elapsed between test and retest measurements was not significantly associated with absolute changes (|estimate| < 0.01 mmHg/day, one-sided p = 0.74) (Fig 3A). Higher proportions of alcoholic patients in the sample were associated with lower absolute changes (estimate = -1.85 mmHg between 0% and 100%, p < 0.001) (Fig 3B). Single-centre studies showed significantly lower absolute changes compared with multicentre studies (estimate = -0.81 mmHg, one-sided p < 0.001) (Fig 3D).

**Fig 3:**
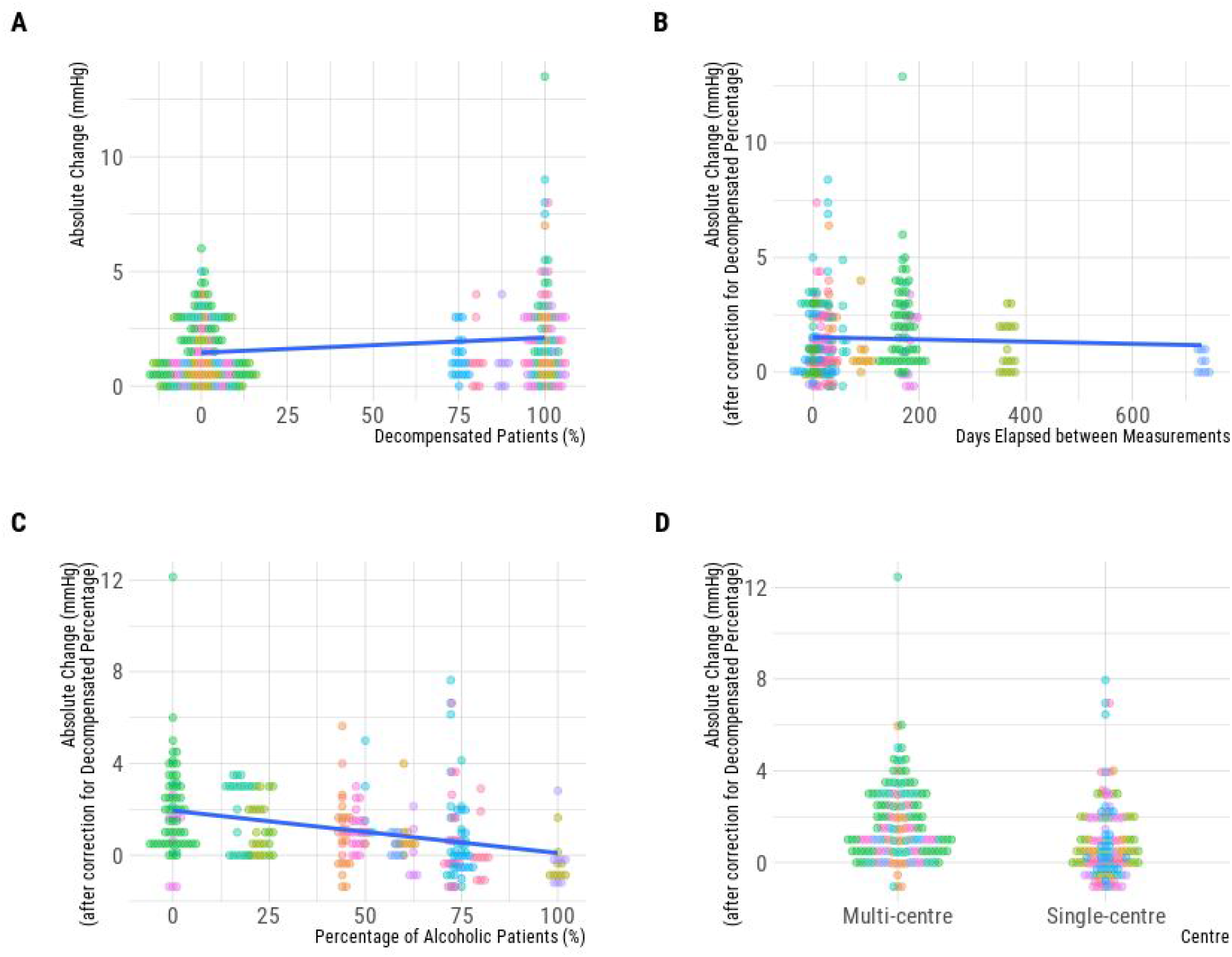
The influence of study characteristics on the absolute change between measurements. Colours represent different studies. Note that because all independent variables are studywide averages, they are the same for all values in each study: for this reason, they are clustered in beeswarm plots around their study mean value. In B, C and D, these changes are shown after correction for the effects of the proportion of decompensated patients: for this reason, some “absolute changes” can be below zero after correction. (A) Absolute change showed a positive association with the percentage of decompensated patients. (B) There was no significant association between the days elapsed between measurements and the absolute changes. (C) Higher percentages of alcoholic patients were significantly associated with lower absolute changes. (D) Single-centre studies showed a significantly lower absolute change between measurements compared to multi-centre studies.

To examine whether these effects are additive, we defined an additional model which included all of the above significant effects, using the same one-sided and two-sided p value thresholds. All effects remained significant in combination: the proportion of compensated cirrhosis patients (estimate = 1.5 mmHg between 0% and 100%, one-sided p < 0.001), the proportion of alcoholic patients (estimate = -1.5 mmHg between 0% and 100%, p < 0.001) and whether the study was conducted at a single centre (estimate = -0.46 mmHg, one-sided p = 0.031).

### Implications for sample size calculations for studies in which HVPG is used as an outcome measure

For assessing the impact of test-retest reliability for studies assessing the effect on HVPG of an intervention, we considered two different scenarios. The first is that of single-arm trials in which the effect of the intervention is assessed without a control group (such as, for example, in early development or proof-of-concept studies). In this scenario, the sample size is selected for a certain size of within-subjects effect. The second scenario would be that of a two-arm parallel randomized trial, in which the goal is to assess the “difference in differences” between two groups. In this scenario, the sample size is selected for a difference of effects between a treated and control group, or to compare two treatments. In both cases, the effect size is defined as the smallest effect of interest, for which the study has greater than a defined level of power to detect effects larger than this value. Since we have shown that reliability metrics are different between studies including only compensated patients *vs* studies including decompensated patients, and it is recommended to study these subpopulations separately^14^, we performed sample size calculations for these two groups of patients.

Further, we considered scenarios in which the effect of the test drug decreasing portal pressure would be homogeneous across study subjects, and scenarios in which the effect would be heterogeneous. An illustration of this concept and further details of these assumptions are shown in supplementary materials S3.

#### a. Single Arm Trials

First, we considered percentage differences for various effect sizes. This addresses the question of the appropriateness of binarising the output variable into treatment response for apparent changes of 10% or 20%. We will consider the case of treatment effects in compensated cirrhosis patients, where the effects are homogeneous (i.e. 100% of patients experience treatment response), as this is a best-case scenario. For no difference, the percentage of patients exhibiting apparent (i.e. measured) changes (i.e. false positives) of 10% or more is 18%, and for 20% changes is 9%. If there is a true change of 1 mmHg following treatment, i.e. 100% of patients experienced treatment effects of this magnitude, the percentage of patients exhibiting apparent changes of 10%+ is 37% and 20%+ is 11%. If there is a true effect of 3mmHg, the percentage of patients exhibiting apparent changes of 10%+ is 80% and 20%+ is 47%. From these figures, it is clear that there are not only a large number of false positives in the case of no effect, but also that there are a large number of false negatives in the case of not only small, but even large effects. The full table is presented in Supplementary Materials S6.

The results of the power analysis show that, for 80% power to detect an effect, true underlying homogeneous differences of require sample sizes (i.e. patients, each of whom is measured before and after treatment), for compensated and decompensated patients respectively, of 27 and 54 for 1 mmHg, 8 and 15 for 2 mmHg; and 5 and 8 for 3 mmHg (Figure 4). Studies in decompensated patients require larger numbers of patients due to the higher degree of measurement error in this group. The full table is presented in Supplementary Materials S7.

**Fig 4:**
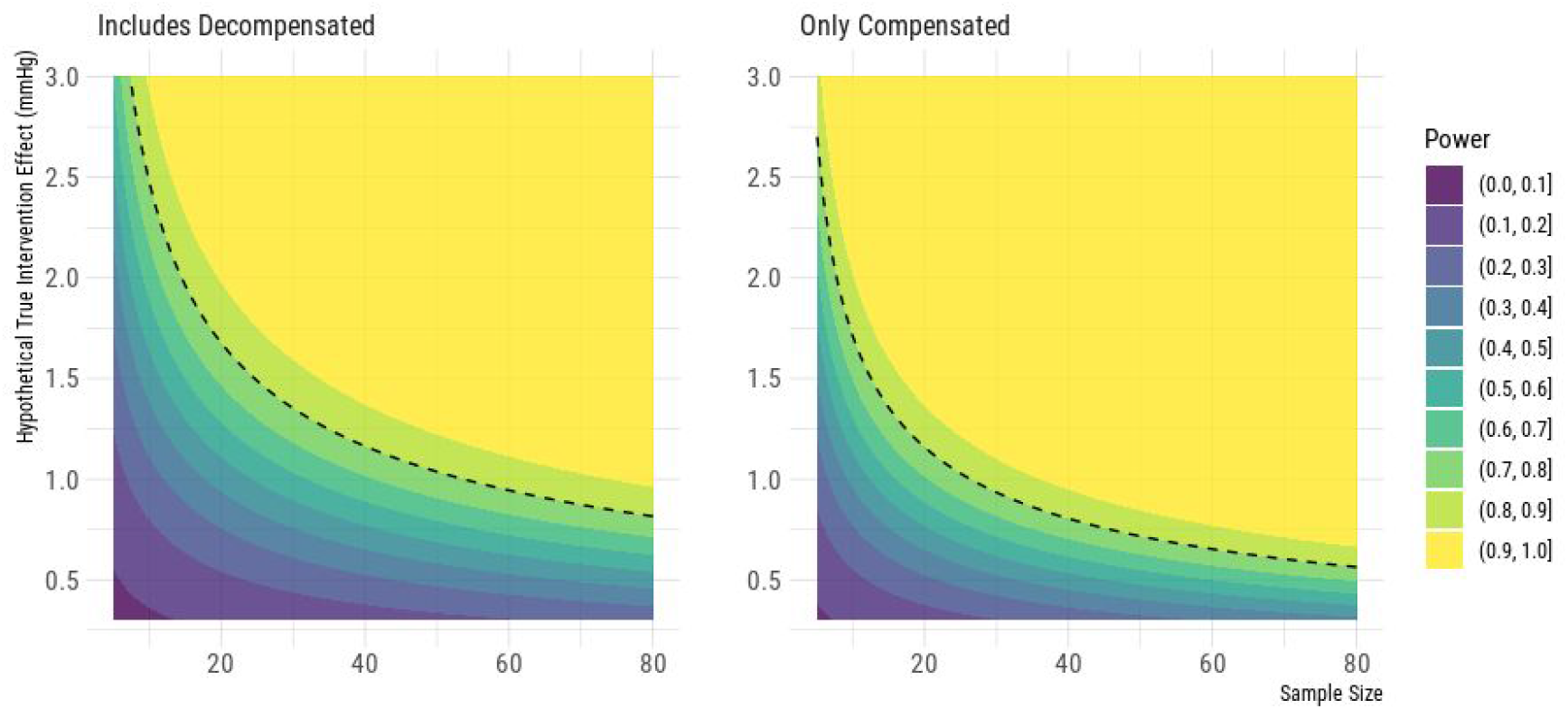
Power contour plots assuming homogeneous effects for different degrees of hypothetical true effect for a within-subject (single-arm) study design of compensated or decompensated patients. The dashed line represents 80% power. By selecting a sample size for which the minimum intervention effect size of interest is adequately powered, a study will have adequate power to detect all potential true intervention effect sizes larger than this value.

#### b. Two-arm parallel randomized trial

Here we performed power analysis to estimate the required sample sizes needed for an RCT comparing two groups under a 1-sided hypothesis test, based on the predicted magnitude of the difference in the change in HVPG in the control and intervention arms for trials studying compensated and decompensated cirrhosis, where the control could be either a placebo (i.e. no change) or a less-effective treatment. For the case of 80% power to detect differences between a placebo arm and an intervention arm, the required sample sizes (i.e. number of patients in each group separately), for compensated and decompensated patients respectively, are 50 and 95 for 1 mmHg, 15 and 30 for 2 mmHg, and 10 and 15 for 3 mmHg (figure 5). The full tables are presented in Supplementary Materials S8.

**Fig 5:**
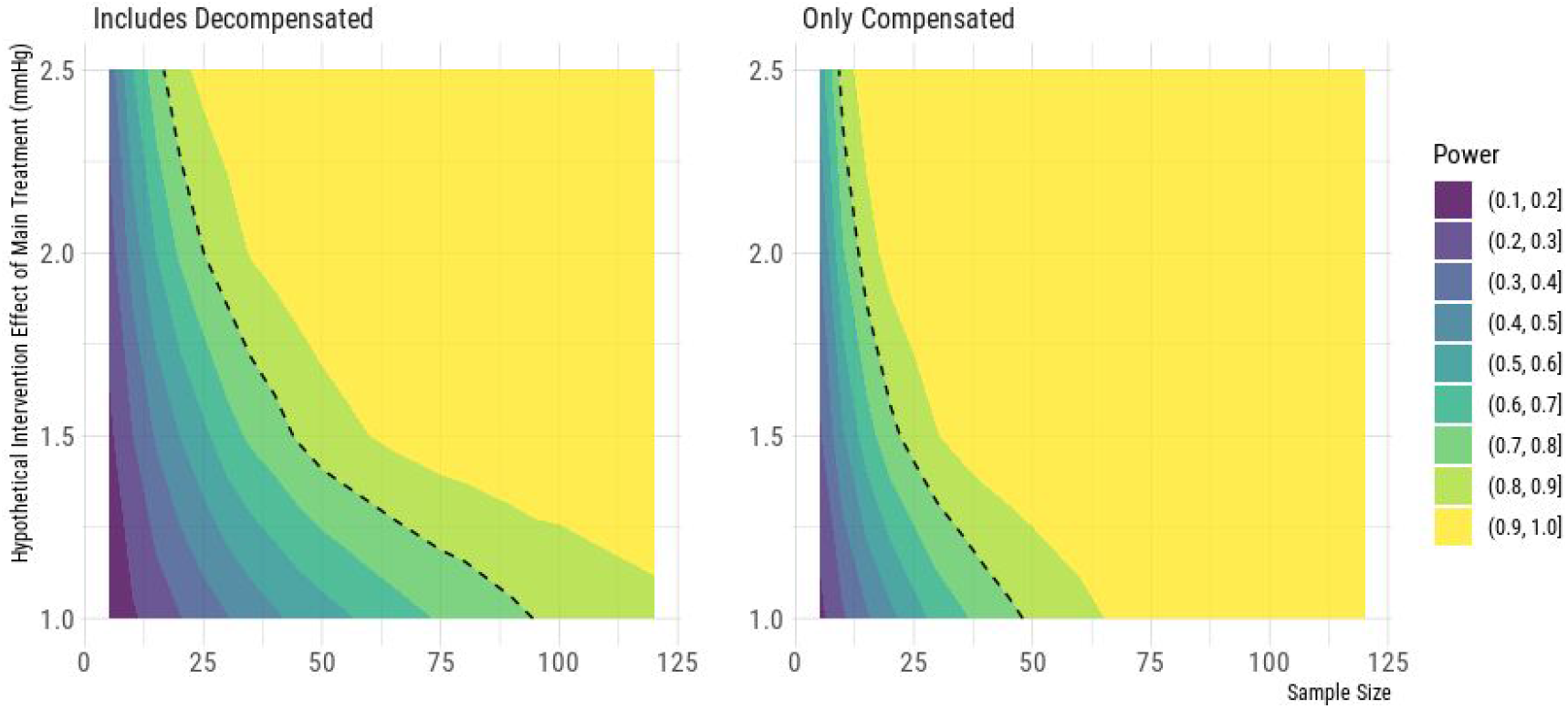
Power contour plots assuming homogeneous effects for different degrees of hypothetical true effect for a two-arm study design comparing a treatment against placebo, for compensated or decompensated patients, under a 1-sided hypothesis test. The dashed line represents 80% power. By selecting a sample size for which the minimum intervention effect size is adequately powered, a study will have adequate power to detect all potential true intervention effect sizes larger than this value.

## Discussion

In this study we show the test-retest reliability and consistency of HVPG, based on repeated HVPG measurements in the context of randomized controlled trials with a placebo or untreated arm. Since this is one of the major contexts of use of HVPG (drug development in portal hypertension), this will facilitate trial design by quantifying the “noise” that investigators might expect when using HVPG as an outcome measurement, and in that way refine trial design. Previous studies have assessed the variability of a single HVPG measurement according to the type of catheter ^22^, intrahepatic heterogeneity of hepatic vein ^45^ or according to the post-hoc reading of permanent tracings by different investigators ^23^. We address here a different research question, which is the reliability and consistency of the measurements using *two different time points* without an active treatment in between, and in the context of RCTs. This specific approach better reflects the specific questions that an investigator faces when planning a RCT based on pre-post HVPG measurements. It also allows a better understanding of the meaning of HVPG change in individual patients which, even if it is not standard of care, has been proposed as a decision rule for therapeutic decisions in patients with cirrhosis and portal hypertension ^46^.

Two major results of the present study are that even if the median change in the placebo groups of these trials was zero (figure 2B), both in trials with compensated and decompensated patients, differences between baseline and follow-up measurements were lower in trials only including compensated patients and trials including decompensated patients. Compensated cirrhosis is a relatively stable condition, with slow changes, and in which portal pressure would be expected to be more stable than in decompensated patients. Patients with decompensated cirrhosis are likely to have more unstable portal pressure, due to disease progression/regression, changes in volemia or repeated bacterial translocation ^47^. Our findings suggest the need of different sample size estimates for trials assessing new drugs for portal hypertension in compensated and decompensated cirrhosis, since the expected noise in HVPG measurements in decompensated patients is higher. Another, somewhat, expected finding, was that the variability in the measurements was higher in multicenter trials than in single center studies, but the magnitude of this effect was low. Finally, studies including higher proportion of alcohol-related cirrhosis patients showed lower variability. Indeed, patients with alcohol-related liver disease considered for these trials are generally abstinent and stable at baseline, and the chances of reaching the second study are higher if this continues to be so during the study period.

Robust evidence shows that, when evaluated at a group level, baseline HVPG or “HVPG response” hold prognostic information in cirrhosis, and this has provided a basis for understanding key elements of the determinants of cirrhosis outcomes, and to develop pharmacological therapies for cirrhosis. The results of this study, however, highlight the difficulties in assessing the HVPG response to a drug *in an individual patient*. In compensated patients, at a group level, within-group differences as small as 1 mmHg can be detected with only ∼30 patients, or between two groups with less than 50 patients per arm. In contrast, at the individual patient-level most studies show that it would be difficult to tell if differences lower than 20% would be related to noise or to true changes related to the drug effect, and this is more so for patients with decompensated cirrhosis (figure 1c). The relative invasiveness and the cost of HVPG limits the possibility of obtaining several measurements at different time points to assess the response to a drug (such as it is done with arterial hypertension, a biomarker with wide physiological variability, where repeated measurements or 24-hr monitoring are used to assess response and escalate therapy in clinical practice ^48^. In the context of trials, it precludes conducting several crossover studies, which is needed to reliably detect individual treatment responses ^49^. These considerations raise caution over approaches to tailor pharmacological therapy according to individual HVPG response or to indicate an escalation of therapy in “non-responders”, and emphasizes the notion that these strategies should be extensively validated in RCTs before being implemented in practice ^50^.It also argues against using the proportion of patients achieving a threshold response to the treatment (or “proportion of responders”) as a clinical relevant outcome, since it is both statistically inefficient and of uncertain clinical interpretation due to the above mentioned considerations (supplementary materials S8) ^51^.

Our study has significant strengths. The study assesses the reliability of HVPG in one of the contexts that it is likely to be used, i.e. randomized controlled trials to compare a new intervention in portal pressure *vs* a standard therapy or a placebo. The range of baseline HVPGs is the range that would be expected to be found in patients with compensated and decompensated cirrhosis, and not in a healthy population that is typically used for test-retest studies in other contexts^24^. Finally, we were able to gather data from 20 studies with 578 HVPG readings in 289 patients, which is a large sample size for a test-retest study, allowing us to derive precise estimates of the reliability and consistency with great precision.

Our analysis has limitations. There might be a study selection bias, since many RCTs in this context have not reported individual patient data (reporting only mean changes). It could be that observation of more test-retest variability might lead to less reporting of individual data. If true, this would lead to an overestimation of reliability in our study. In addition, especially in long term studies, there might be a patient selection bias, since those patients that reached the second measurement are generally those that did not have a major clinical event and, therefore, these patients might have a more stable portal pressure. However, the reported attrition in these trials was very low, limiting the impact of this potential bias. In this regard, we were only able to retrieve 92% of all measurements theoretically reported in the different studies (table 1). This might relate to lack of reporting of patients not reaching the second HVPG measurement, or to the fact that two patients with the exact same trajectory would have overlapping lines or dots. The latter would have a minor impact on reliability metrics, since it means that these patients did not have more extreme values than the ones analysed here. Finally, we had limited capacity to assess the factors that might determine the reliability of HVPG in clinical trials, since we could only assess study-level characteristics.

In summary, using data from untreated patients in clinical trials, we show that HVPG exhibits good reliability and consistency, which is higher in compensated than decompensated patients. Using the results of this analysis, we performed simulation-based power analyses for sample size calculation for trials in which HVPG is to be used as a readout for the evaluation of new drugs. Studies in decompensated patients would need higher sample sizes than studies including only compensated patients, since the repeatability of HVPG is lower in the former. This study also raises caution to the proposal of switching therapies based on reaching or not reaching a defined threshold of HVPG response, especially in decompensated patients.

## Data Availability

All raw data, analysis code and additional figures can be found at the following link: https://github.com/mathesong/HVPG_TRT

https://github.com/mathesong/HVPG_TRT

## Supplementary Materials 1

### Search Strategy

We performed a search on PUBMED and Ovid MEDLINE from database inception through to January 2020. After removing duplicates, we screened potential eligible records, initially at title and abstract and then at full text level. Search and selection of the studies was designed by JS, WB and JGA with the collaboration of the University of Alberta Faculty of Medicine librarians. Final assessment of the articles was conducted by WB and JS. Any discrepancies were resolved through consulting JGA.

### Search Synthax

MEDLINE (via PubMed) 1970 to Jan 2020

#1 HVPG

#2 Hepatic venous pressure gradient

#3 Hepatic pressure gradient

#4 Portal pressure

#5 Portal haemodynamic*

#6 Portal hemodynamic*

#7 porto-systemic*

#8 portosystemic*

#9 #1 Or #2 OR #3 OR #4 OR #5 OR #6 OR #7 OR #8

#10 Placebo

#11 Cirrhosis

#12 Chronic liver disease

#13 #11 OR #12

#14 #9 AND #10 AND #13

#15 “Hypertension Portal”[mesh]

#16 “Liver cirrhosis/ complications and drug therapy[mesh]

#17 “Portal pressure/drug effects”[mesh]

#18 “Portal system/drug effects”[mesh]

#19 #17 OR #18

#20 #15 AND #16 AND #19

An additional search was performed by reviewing recent reviews on novel treatments for portal hypertension. We did an additional search in Google Images with the following terms: HVPG AND (Placebo OR spaghetti plot OR pre-post). These strategies identified 4 additional trials not picked up in the structured search.

### Flow diagram showing study selection

**Supplementary Figure 1.**
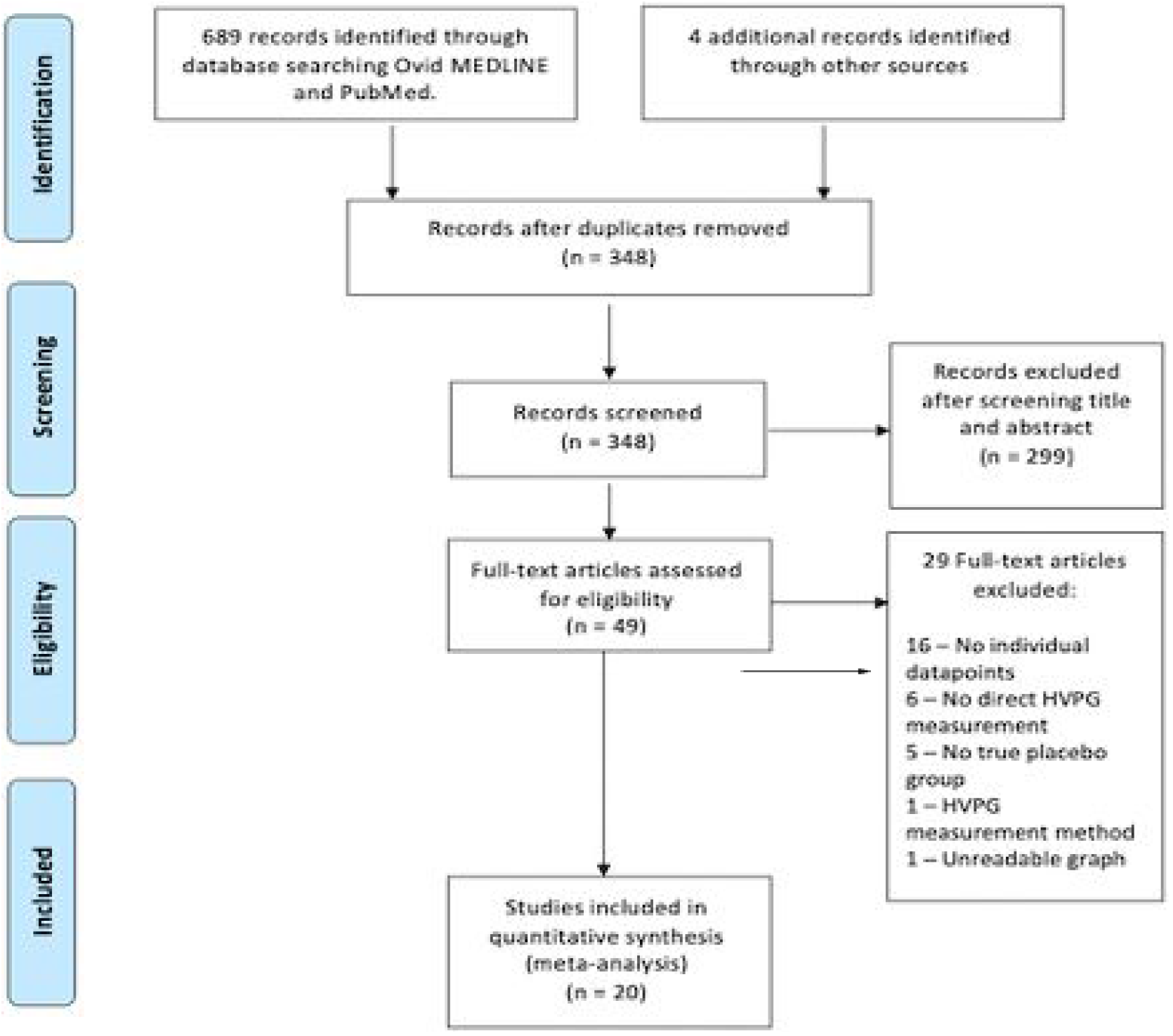
Flow diagram.

## Data Extraction

Baseline and follow-up HVPG measurements in the control group were extracted after digitizing the plots showing individual patient HVPG data. Two observers (WB and MAK) independently extracted the values of the baseline and follow-up HVPG by creating a digital grid over the published figures. Any discrepancies were resolved by consensus involving a third reviewer (JGA). For assessing the characteristics of the source studies two reviewers (WB and JS) independently extracted the data shown in table 1. Any discrepancies were resolved through consensus involving a third reviewer (JGA).

## Examples of readings of HVPG plots

In this partial plot, in the x-axis the authors present the baseline HVPG. In the y-axis, the change at day 28. we show how the overlying grid allows to extract a value of 19.5 mmHg for the baseline, and a change of -13.5 mmHg, which means that the follow-up value was 6 mmHg.

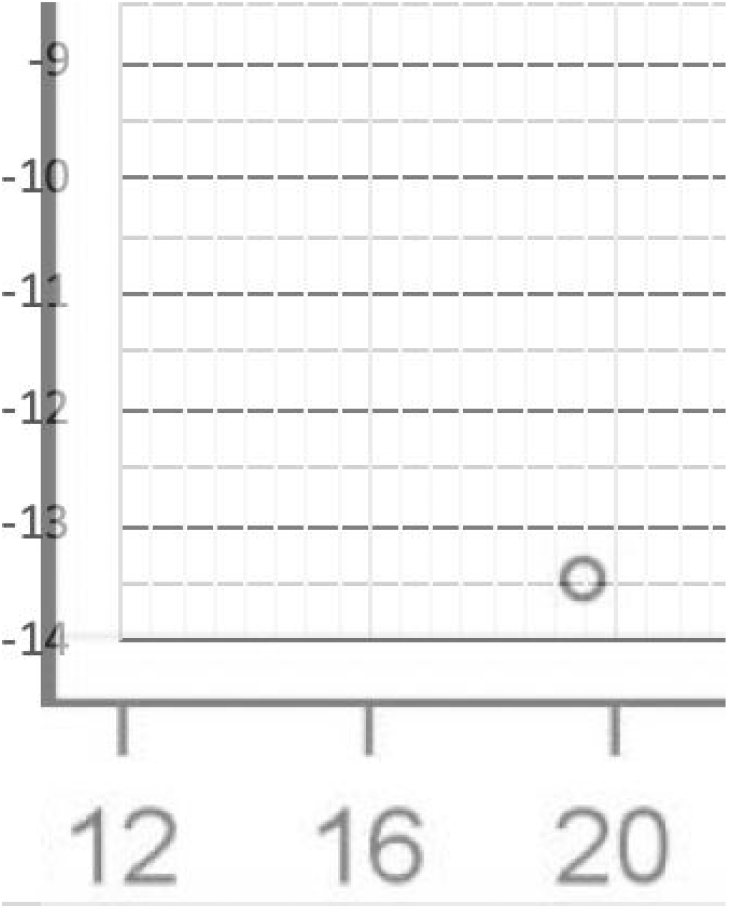

The following partial graph illustrates the application of this methodology to a “spaghetti plot”. The case starting with an HVPG of 26 mmHg ends up with a follow-up HVPG of 29 mmHg.

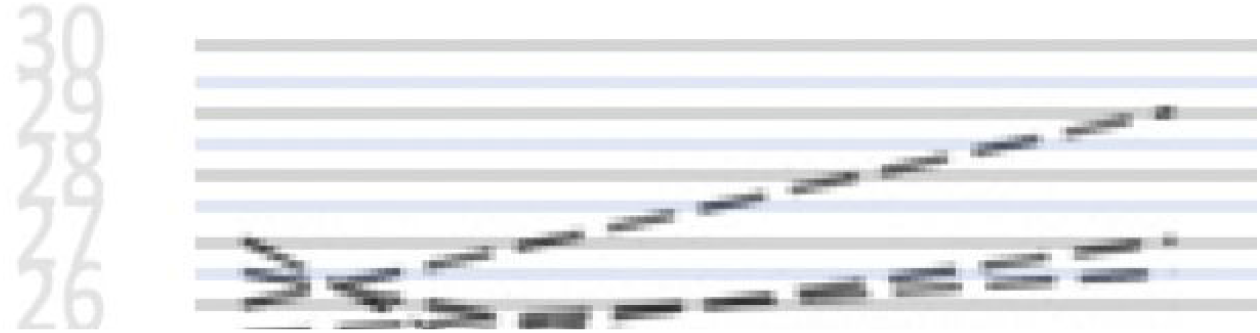

## Supplementary materials 2

### Illustration of the concept of intraclass correlation coefficient

The ICC is defined as the fraction of the total variance (the sum of the true and the error variance) which is not attributable to error (i.e. within-individual changes). Here, we visualise the degree of variability originating from both components and the total to give readers a feeling for the meaning of the ICC. The distributions show the implied likelihood of each value, given the true mean value (dashed line). True inter-individual variance is in green (i.e., between-individual variability of the underlying ‘true’ values), measurement error variance is in maroon (i.e. within-individual variability of the measured values), and the total (i.e. measured: the sum of both true and error) variance is in black for different intraclass correlation coefficients (ICC). It follows that with ICC < 0.5, variance around each individual’s true value from measurement to measurement will be greater than the amount of true inter-individual variability around the true mean.

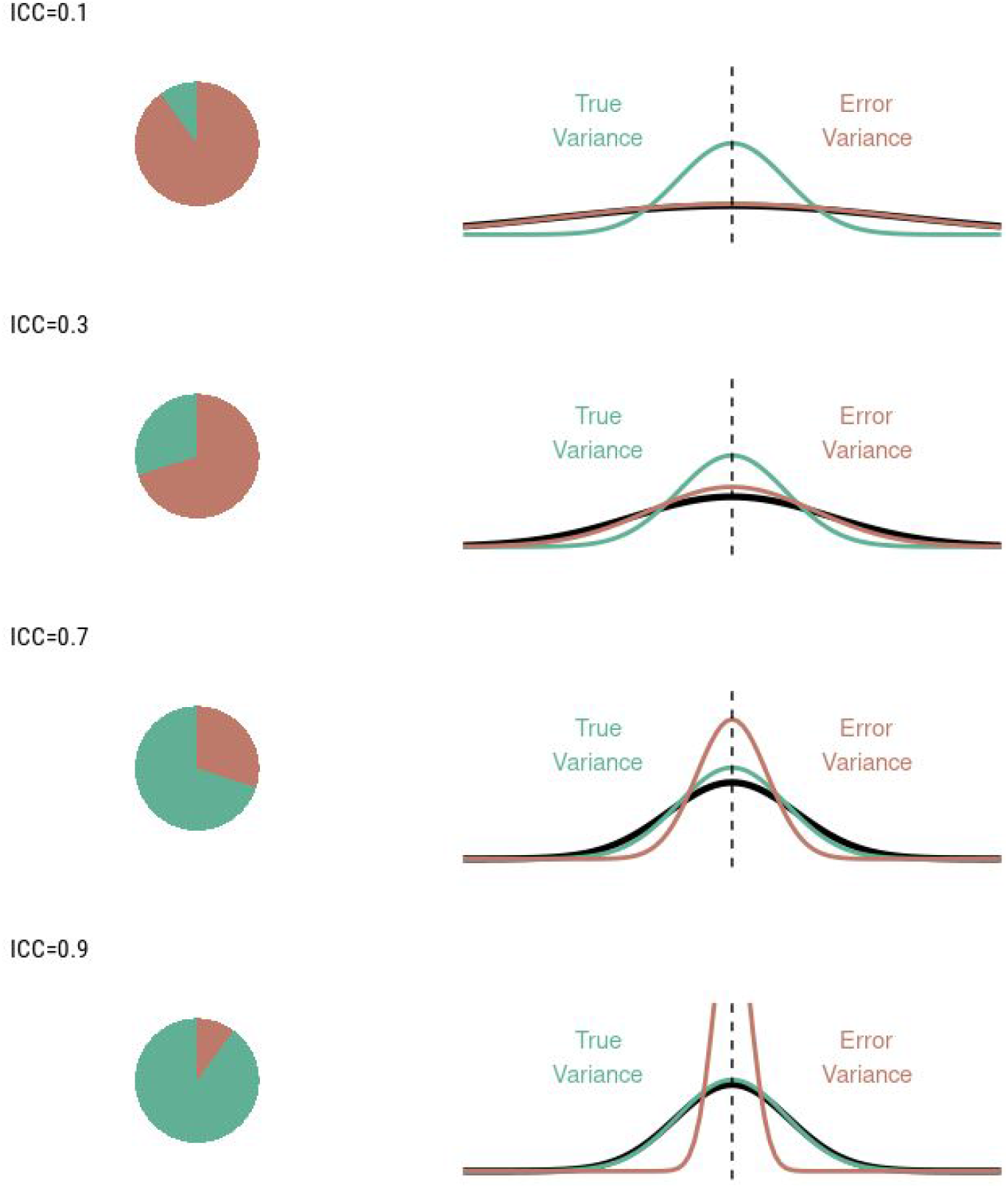

### Supplementary materials 3

Study-level ICCs and SDDs were calculated using the R packages *relfeas*^*1*^, *psych*^*2*^ and *agRee*^*3*^. ICCs and SDDs were statistically compared between all studies of compensated patients and those including decompensated patients using a case-resampling bootstrap procedure. To assess the role of the study characteristics on the degree of measurement error of HVPG, we assessed their influence on the within-patient absolute (i.e. unsigned) change. We made use of permutation tests using the R packages *perm*^*4*^ and *permuco*^*5*^ on account of the high degree of skew (=2.14) in the distribution of absolute change to mitigate the effect of influential points. When assessing the signed changes, which were more normally distributed (skew = -0.73), we made use of linear mixed effects models, using the source study as a random intercept in order to account for the hierarchical, clustered, nature of the data with measurements coming from different studies. In all cases, tests were performed after correction for the percentage of the sample comprised of decompensated patients when examining the total sample.

Power calculations were performed using the *pwr* R package for within-individual differences (one-arm trials)^6^. For comparing differences between two groups (two-arm parallel trials), power calculations were performed by simulating data (10,000 datasets for each configuration) with the same measurement characteristics estimated from the test-retest data. Hence, “true” values were created around the overall mean with the true variance (i.e. the ICC multiplied with the total variance), and measurement error was created using the within-subject coefficient of variation. We also examined the effect of potential heterogeneity in treatment effects: all power analyses were performed both for homogeneous underlying true effects (i.e. assuming every individual changed by the same amount before the addition of measurement error), as well as for heterogeneous underlying true effects (i.e. some individuals experience greater effects than others). For the heterogeneous effects, we applied a coefficient of variation around the differences of 50%. This implies that 2.3% of patients would experience a true worsening of symptoms following treatment, while a different 2.3% would experience an improvement of over double the mean effect. This is further illustrated in the figure

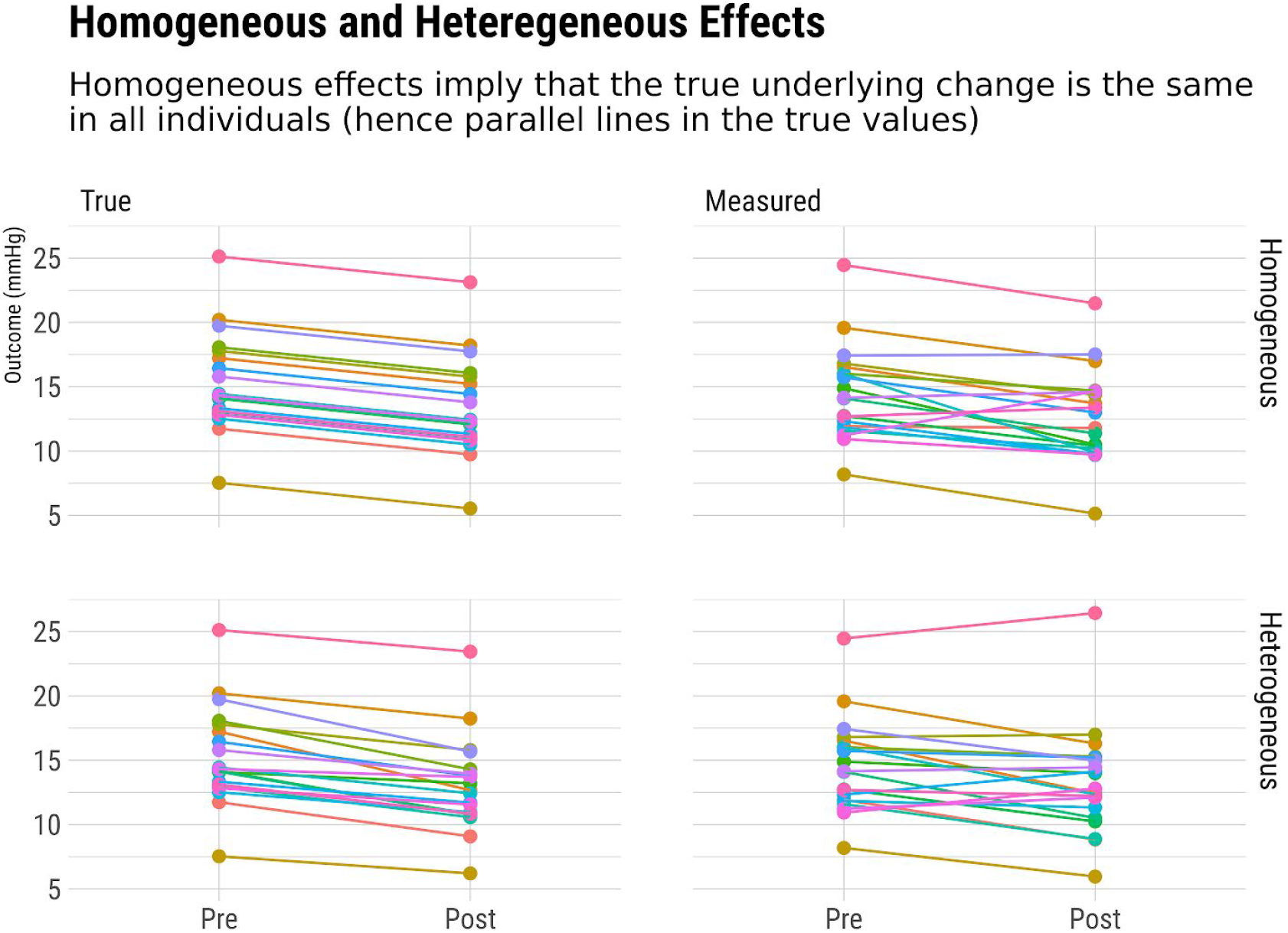

*Figure: The figure above shows theoretical underlying true values (left), and measured values including measurement error (right). The true change from before to after the intervention can either be homogeneous (i*.*e. everyone has exactly the same effect, upper panels), or heterogeneous (i*.*e. some individuals experience greater effects of the intervention than others, lower panels). For heterogeneous effects, we applied a coefficient of variation of the intervention effect of 50%, which implies that 2*.*3% of the sample would experience a true worsening, and 2*.*3% of the sample would experience over double the effect of the mean. In all cases above, the mean change is a decrease of 2 mmHg. Importantly, due to measurement error, it will always appear as if some individuals experience greater treatment effects than others, even with a homogeneous change in portal pressure. However, if the effects of treatment are truly heterogeneous, then the variance will be greater. With greater variance, the standardised effect size will be smaller, and therefore the power will be less to detect an effect of the same absolute magnitude (e*.*g. 2mmHg). As shown in the sample size calculations, if a heterogeneous effect of the drug is assumed, the sample size calculation to detect a given change in HVPG will be slightly higher*.

## Supplementary data 4

### Summary of study characteristics

A total of 578 HVPG measurements in 289 patients were identified from the plots, out of the reported 313 patients included in the control groups of these studies (92% retrieval rate). Most studies had placebo arms and only in 3 studies the control arm was composed of untreated patients. Only one study reported that HVPG measurements were conducted with a straight “wedged” catheter^1^. One additional study reported measurements with both balloon and wedged catheter in an undefined proportion^2^. The latter study was considered as a “balloon catheter study” for further analysis.

The study by Pomier-Layrargues et al^3^ included three sequential measurements of HVPG. The first was performed just after an episode of acute variceal bleed, the second at ten days after the bleed, and the third at 6 months. Only the values at 10 days and at 6 months were used for test-retest reliability since values obtained immediately after variceal bleeding were considered to be unstable. Indeed, there was a major drop in HVPG between the first and the second measurement.

In the studies that reported the inclusion of decompensated patients, and in which the results of compensated and decompensated patients could not be individualized, the proportion of decompensated patients was over 50% (table 1) and were considered within the group of studies assessing “decompensated” studies.

## Abbreviations

HVPG: Hepatic venous pressure gradient
CV: coefficient of variation
WSCV: Within-subject coefficient of variation
ICC: intraclass correlation coefficient
SDD: smallest detectable difference.
Change SD: the standard deviation of the signed change between measurements

## Supplementary materials 5

**Table:** Metrics of reliability and consistency of the 20 included studies, divided by whether they included decompensated patients or not. A detailed interpretation of these measurements is provided in Matheson, PeerJ. 2019; 7: e6918.

| Study | n | Decompensated (%) | Alcoholic (%) | Mean Days Elapsed | Mean | CV | WSCV | ICC | SDD | Change SD |
| --- | --- | --- | --- | --- | --- | --- | --- | --- | --- | --- |
| **Only Compensated** |  |  |  |  |  |  |  |  |  |  |
| Abraldes 2008 (C) | 16 | 0.0 | 44.0 | 30.00 | 18.47 | 0.16 | 0.07 | 0.81 | 3.65 | 1.94 |
| Albillos 1995 | 20 | 0.0 | 60.0 | 90.00 | 19.60 | 0.20 | 0.05 | 0.94 | 2.74 | 1.46 |
| Berzigotti 2010 | 4 | 0.0 | 50.0 | 16.00 | 18.62 | 0.12 | 0.03 | 0.96 | 1.55 | 1.06 |
| Debernardi 2007 | 34 | 0.0 | 21.7 | 365.00 | 14.59 | 0.12 | 0.07 | 0.64 | 3.01 | 1.40 |
| Fukada 2014 | 16 | 0.0 | 25.0 | 0.04 | 17.28 | 0.24 | 0.07 | 0.93 | 3.19 | 1.71 |
| Garcia-Tsao 2020 (C) | 100 | 0.0 | 0.0 | 168.00 | 16.63 | 0.24 | 0.10 | 0.82 | 4.75 | 2.44 |
| Hidaka 2011 | 38 | 0.0 | 16.7 | 0.01 | 14.75 | 0.25 | 0.12 | 0.79 | 4.80 | 2.37 |
| Lebrec 2012 | 12 | 0.0 | 50.0 | 0.04 | 18.04 | 0.17 | 0.10 | 0.66 | 5.01 | 2.54 |
| Merkel 2004 | 18 | 0.0 | 57.7 | 730.00 | 12.39 | 0.10 | 0.04 | 0.87 | 1.22 | 0.62 |
| Reverter 2015 | 42 | 0.0 | 47.6 | 15.00 | 15.76 | 0.28 | 0.06 | 0.95 | 2.76 | 1.34 |
| **Includes Decompensated** |  |  |  |  |  |  |  |  |  |  |
| Abraldes 2008 (D) | 36 | 100.0 | 44.0 | 30.00 | 20.28 | 0.22 | 0.10 | 0.81 | 5.43 | 2.77 |
| Blei 1987 | 18 | 100.0 | 100.0 | 0.04 | 15.06 | 0.31 | 0.06 | 0.97 | 2.49 | 1.06 |
| Garcia-Tsao 2020 (D) | 14 | 100.0 | 0.0 | 168.00 | 16.32 | 0.29 | 0.27 | 0.26 | 12.17 | 5.06 |
| Jayakumar 2013 | 16 | 100.0 | 75.0 | 56.00 | 22.72 | 0.14 | 0.09 | 0.62 | 5.42 | 2.96 |
| Kimer 2017 | 36 | 100.0 | 72.2 | 28.00 | 16.18 | 0.28 | 0.17 | 0.63 | 7.62 | 3.98 |
| McCormick 1992 | 40 | 75.0 | 75.0 | 0.01 | 17.44 | 0.25 | 0.06 | 0.94 | 3.04 | 1.56 |
| Moller 2000 | 16 | 87.5 | 100.0 | 0.02 | 15.38 | 0.35 | 0.07 | 0.96 | 3.10 | 1.67 |
| Pomier 1987 | 12 | 100.0 | 62.5 | 172.50 | 15.42 | 0.36 | 0.09 | 0.95 | 3.71 | 1.26 |
| Pozzi 2005 | 18 | 100.0 | 0.0 | 182.50 | 16.11 | 0.22 | 0.11 | 0.78 | 4.89 | 2.24 |
| Schepke 2001 | 36 | 100.0 | 72.5 | 7.00 | 18.25 | 0.21 | 0.12 | 0.69 | 6.01 | 3.15 |
| Schwarzer 2017 | 20 | 80.0 | 80.0 | 28.00 | 20.50 | 0.24 | 0.06 | 0.94 | 3.39 | 1.78 |
| Spahr 2007 | 16 | 75.0 | 62.5 | 90.00 | 17.69 | 0.17 | 0.16 | 0.14 | 7.64 | 3.95 |

n: number of observations (includes baseline and follow-up HVPG). CV: coefficient of variation. WSCV: Within-subject coefficient of variation. ICC: intraclass correlation coefficient. SDD: smallest detectable difference. Change SD: the standard deviation of the signed change values between measurements, helpful for power analysis for longitudinal studies.

## Supplementary material 6

Table: Apparent percentage of responders, defined as 10% or 20% decrease in portal pressure, according to the true underlying change in HVPG (data modelled using the reliability metrics obtained in the present study). As shown in the table, even in the case of a true zero homogeneous effect, 18% of a group of compensated patients, and 25% of decompensated patients would show an apparent 10% decrease in portal pressure.

| Difference | Effects | True Worsening (%) | Apparent Worsening (%) | True 10%+ Improvement (%) | Apparent 10%+ Improvement (%) | True 20%+ Improvement (%) | Apparent 20%+ Improvement (%) |
| --- | --- | --- | --- | --- | --- | --- | --- |
| Includes Decompensated |  |  |  |  |  |  |  |
| 0.0 | Homogeneous | 0.0 | 50.0 | 0.0 | 25.4 | 0.0 | 9.1 |
| 0.0 | Heterogeneous | 0.0 | 50.0 | 0.0 | 25.4 | 0.0 | 9.1 |
| 0.5 | Homogeneous | 0.0 | 42.8 | 0.2 | 31.8 | 0.0 | 12.7 |
| 0.5 | Heterogeneous | 2.3 | 42.8 | 0.6 | 31.9 | 0.0 | 12.8 |
| 1.0 | Homogeneous | 0.0 | 35.8 | 3.9 | 38.8 | 0.2 | 17.1 |
| 1.0 | Heterogeneous | 2.3 | 36.0 | 12.7 | 39.0 | 0.6 | 17.5 |
| 1.5 | Homogeneous | 0.0 | 29.2 | 27.8 | 46.3 | 1.0 | 22.3 |
| 1.5 | Heterogeneous | 2.3 | 29.9 | 38.6 | 46.3 | 3.9 | 23.2 |
| 2.0 | Homogeneous | 0.0 | 23.3 | 72.0 | 53.8 | 3.9 | 28.4 |
| 2.0 | Heterogeneous | 2.3 | 24.7 | 59.1 | 53.5 | 12.7 | 29.7 |
| 2.5 | Homogeneous | 0.0 | 18.1 | 96.0 | 61.2 | 12.0 | 35.1 |
| 2.5 | Heterogeneous | 2.3 | 20.4 | 71.5 | 60.1 | 25.4 | 36.5 |
| 3.0 | Homogeneous | 0.0 | 13.7 | 99.8 | 68.2 | 27.9 | 42.4 |
| 3.0 | Heterogeneous | 2.3 | 16.9 | 78.8 | 65.9 | 38.6 | 43.4 |
| Only Compensated |  |  |  |  |  |  |  |
| 0.0 | Homogeneous | 0.0 | 50.0 | 0.0 | 17.7 | 0.0 | 3.5 |
| 0.0 | Heterogeneous | 0.0 | 50.0 | 0.0 | 17.7 | 0.0 | 3.5 |
| 0.5 | Homogeneous | 0.0 | 38.7 | 0.2 | 26.3 | 0.0 | 6.4 |
| 0.5 | Heterogeneous | 2.3 | 38.9 | 0.7 | 26.5 | 0.0 | 6.6 |
| 1.0 | Homogeneous | 0.0 | 28.3 | 5.7 | 36.7 | 0.2 | 10.8 |
| 1.0 | Heterogeneous | 2.3 | 29.1 | 17.6 | 37.2 | 0.7 | 11.7 |
| 1.5 | Homogeneous | 0.0 | 19.5 | 41.7 | 48.2 | 1.2 | 17.1 |
| 1.5 | Heterogeneous | 2.3 | 21.5 | 46.3 | 48.4 | 5.7 | 19.2 |
| 2.0 | Homogeneous | 0.0 | 12.6 | 87.7 | 59.9 | 5.7 | 25.4 |
| 2.0 | Heterogeneous | 2.3 | 16.0 | 65.4 | 58.5 | 17.6 | 28.3 |
| 2.5 | Homogeneous | 0.0 | 7.6 | 99.4 | 70.7 | 18.5 | 35.4 |
| 2.5 | Heterogeneous | 2.3 | 12.2 | 76.1 | 66.9 | 32.6 | 38.0 |
| 3.0 | Homogeneous | 0.0 | 4.3 | 100.0 | 79.9 | 41.7 | 46.5 |
| 3.0 | Heterogeneous | 2.3 | 9.6 | 82.2 | 73.5 | 46.3 | 47.3 |

## Supplementary material 7

Power calculations to determine sample sizes for a single arm trial, according to whether the study would include decompensated/compensated patients, the true effect size, and whether true effects are homogeneous or heterogeneous.

| Difference | Effects | 80% Power | 90% Power |
| --- | --- | --- | --- |
| **Includes Decompensated** |  |  |  |
| 1.0 | Homogeneous Effects | 54 | 74 |
| 1.0 | Heterogeneous Effects | 56 | 76 |
| 1.5 | Homogeneous Effects | 25 | 34 |
| 1.5 | Heterogeneous Effects | 27 | 36 |
| 2.0 | Homogeneous Effects | 15 | 20 |
| 2.0 | Heterogeneous Effects | 17 | 22 |
| 2.5 | Homogeneous Effects | 10 | 14 |
| 2.5 | Heterogeneous Effects | 12 | 16 |
| 3.0 | Homogeneous Effects | 8 | 10 |
| 3.0 | Heterogeneous Effects | 9 | 12 |
| **Only Compensated** |  |  |  |
| 1.0 | Homogeneous Effects | 27 | 36 |
| 1.0 | Heterogeneous Effects | 28 | 39 |
| 1.5 | Homogeneous Effects | 13 | 17 |
| 1.5 | Heterogeneous Effects | 15 | 19 |
| 2.0 | Homogeneous Effects | 8 | 11 |
| 2.0 | Heterogeneous Effects | 10 | 13 |
| 2.5 | Homogeneous Effects | 6 | 8 |
| 2.5 | Heterogeneous Effects | 8 | 10 |
| 3.0 | Homogeneous Effects | 5 | 6 |
| 3.0 | Heterogeneous Effects | 6 | 8 |

## Supplementary material 8

Power calculations to determine sample sizes for a two-arm parallel randomized trial under a one-sided hypothesis test, according to whether the study would include decompensated/compensated patients, the true difference between the treatment and the control arm, and whether true effects are homogeneous or heterogeneous

| Difference (mmHg) | Intervention 1 Effect (mmHg) | Intervention 2 Effect (mmHg) | Effects | 80% Power | 90% Power |
| --- | --- | --- | --- | --- | --- |
| Includes Decompensated |  |  |  |  |  |
| 3 | 3 | 0 | Homogeneous Effects | 15 | 20 |
| 3 | 3 | 0 | Heterogeneous Effects | 15 | 20 |
| 2.5 | 3 | 0.5 | Homogeneous Effects | 20 | 25 |
| 2.5 | 3 | 0.5 | Heterogeneous Effects | 20 | 30 |
| 2.5 | 2.5 | 0 | Homogeneous Effects | 20 | 25 |
| 2.5 | 2.5 | 0 | Heterogeneous Effects | 20 | 25 |
| 2 | 3 | 1 | Homogeneous Effects | 30 | 35 |
| 2 | 3 | 1 | Heterogeneous Effects | 30 | 40 |
| 2 | 2.5 | 0.5 | Homogeneous Effects | 25 | 35 |
| 2 | 2.5 | 0.5 | Heterogeneous Effects | 30 | 40 |
| 2 | 2 | 0 | Homogeneous Effects | 30 | 35 |
| 2 | 2 | 0 | Heterogeneous Effects | 30 | 40 |
| 1.5 | 3 | 1.5 | Homogeneous Effects | 45 | 65 |
| 1.5 | 3 | 1.5 | Heterogeneous Effects | 55 | 75 |
| 1.5 | 2.5 | 1 | Homogeneous Effects | 45 | 60 |
| 1.5 | 2.5 | 1 | Heterogeneous Effects | 50 | 70 |
| 1.5 | 2 | 0.5 | Homogeneous Effects | 45 | 60 |
| 1.5 | 2 | 0.5 | Heterogeneous Effects | 50 | 65 |
| 1.5 | 1.5 | 0 | Homogeneous Effects | 45 | 60 |
| 1.5 | 1.5 | 0 | Heterogeneous Effects | 50 | 60 |
| 1 | 3 | 2 | Homogeneous Effects | 100 | 140 |
| 1 | 3 | 2 | Heterogeneous Effects | 120 | 170 |
| 1 | 2.5 | 1.5 | Homogeneous Effects | 100 | 140 |
| 1 | 2.5 | 1.5 | Heterogeneous Effects | 110 | 150 |
| 1 | 2 | 1 | Homogeneous Effects | 100 | 140 |
| 1 | 2 | 1 | Heterogeneous Effects | 110 | 150 |
| 1 | 1.5 | 0.5 | Homogeneous Effects | 100 | 140 |
| 1 | 1.5 | 0.5 | Heterogeneous Effects | 110 | 140 |
| 1 | 1 | 0 | Homogeneous Effects | 95 | 140 |
| 1 | 1 | 0 | Heterogeneous Effects | 100 | 140 |
| Only Compensated |  |  |  |  |  |
| 3 | 3 | 0 | Homogeneous Effects | 10 | 10 |
| 3 | 3 | 0 | Heterogeneous Effects | 10 | 15 |
| 2.5 | 3 | 0.5 | Homogeneous Effects | 10 | 15 |
| 2.5 | 3 | 0.5 | Heterogeneous Effects | 15 | 15 |
| 2.5 | 2.5 | 0 | Homogeneous Effects | 10 | 15 |
| 2.5 | 2.5 | 0 | Heterogeneous Effects | 15 | 15 |
| 2 | 3 | 1 | Homogeneous Effects | 15 | 20 |
| 2 | 3 | 1 | Heterogeneous Effects | 20 | 25 |
| 2 | 2.5 | 0.5 | Homogeneous Effects | 15 | 20 |
| 2 | 2.5 | 0.5 | Heterogeneous Effects | 20 | 25 |
| 2 | 2 | 0 | Homogeneous Effects | 15 | 20 |
| 2 | 2 | 0 | Heterogeneous Effects | 15 | 20 |
| 1.5 | 3 | 1.5 | Homogeneous Effects | 25 | 35 |
| 1.5 | 3 | 1.5 | Heterogeneous Effects | 30 | 45 |
| 1.5 | 2.5 | 1 | Homogeneous Effects | 25 | 30 |
| 1.5 | 2.5 | 1 | Heterogeneous Effects | 30 | 40 |
| 1.5 | 2 | 0.5 | Homogeneous Effects | 25 | 35 |
| 1.5 | 2 | 0.5 | Heterogeneous Effects | 25 | 35 |
| 1.5 | 1.5 | 0 | Homogeneous Effects | 25 | 35 |
| 1.5 | 1.5 | 0 | Heterogeneous Effects | 25 | 35 |
| 1 | 3 | 2 | Homogeneous Effects | 50 | 65 |
| 1 | 3 | 2 | Heterogeneous Effects | 70 | 95 |
| 1 | 2.5 | 1.5 | Homogeneous Effects | 50 | 70 |
| 1 | 2.5 | 1.5 | Heterogeneous Effects | 60 | 85 |
| 1 | 2 | 1 | Homogeneous Effects | 50 | 70 |
| 1 | 2 | 1 | Heterogeneous Effects | 60 | 80 |
| 1 | 1.5 | 0.5 | Homogeneous Effects | 50 | 70 |
| 1 | 1.5 | 0.5 | Heterogeneous Effects | 55 | 75 |
| 1 | 1 | 0 | Homogeneous Effects | 50 | 70 |
| 1 | 1 | 0 | Heterogeneous Effects | 50 | 70 |

